# High Perforation Rates in Jejunal Diverticulitis: A Single-Center Retrospective Review

**DOI:** 10.64898/2026.04.05.26350210

**Authors:** Natalie Florescu, Evelyn C. Thomas, Anthony Charles, Alia Aunchman, Gary An

**Author notes:** Corresponding Author: Natalie Florescu.

## Abstract

**Background:** Jejunal diverticulitis is an uncommon but increasingly recognized cause of acute abdomen. It can present with a range of CT findings, including peridiverticular inflammation, bowel wall thickening, and fecalized small bowel content, with perforation or abscess occurring as complications in roughly 6% of cases. Case reports note varied presentations with jejunal and ileal involvement, treatment ranging from nonoperative management with antibiotics to urgent surgical intervention. Though rare, small bowel diverticulitis, particularly involving the jejunum, can result in significant morbidity, including peritonitis and sepsis, requiring heightened clinical suspicion in elderly or immunocompromised patients.

**Methods:** We conducted a single center retrospective review of patients diagnosed with jejunal diverticulitis in a single academic center’s Emergency General Surgery registry between December 2017 and December 2024. Of 42 patients initially identified, 34 had confirmed diagnoses on chart review. Data abstracted included age, sex, imaging modality, presence of perforation, serial physical exams, lab values (CBC, lactate), ICU admission, length of stay (LOS), antibiotic duration, operative status and timing, distance of residence from our institution, disposition after index admission, and readmission within one year.

**Results:** Of the 34 confirmed cases, 24 (71%) were perforated: 2 presented with small bowel obstruction, 16 with abscesses and/or contained perforations, and 1 with both. 19 of the 24 perforated patients required operative intervention: 9 proceeded directly to the OR, 3 on hospital day one, and 2 as late as hospital day six. Among non-operative patients treated with antibiotics alone, the average LOS was 6 days (range: 2–23). Two patients were readmitted within one year: neither had undergone surgery during their index admission and neither were related to their index admission. Overall, three patients died: two during the index admission (both perforated and operated on) and one on readmission.

**Conclusion:** Compared to the 6% complication rate reported in prior literature, our series demonstrates a notably higher rate of perforation (71%) among patients diagnosed with jejunal diverticulitis. Operative intervention was common, though a subset of patients was successfully managed non-operatively with antibiotics. Mortality was limited to patients with significant comorbidities and complex presentations. These findings underscore the heterogeneity in presentation and outcomes and highlight the need for a standardized approach. Development of practice guidelines incorporating clinical, radiographic, and laboratory parameters may improve diagnostic accuracy and guide timely, evidence-based management of this rare but serious condition.

## Introduction

Diverticula are mucosal outpouchings that occur at weakened areas of the intestinal wall and are thought to result from bowel dyskinesia, increased intraluminal pressure, or areas with greater vasa recta distribution (1). Diverticula can become infected and inflamed, causing diverticulitis (2). Case reports have shown secondary causes to include enteroliths, stromal tumors, and Nuck cysts (1, 3-5). Jejunal diverticulosis is particularly challenging to diagnose and manage, as contained perforations frequently present with subtle clinical and radiographic findings. While the duodenum is traditionally cited as the most common site of small bowel diverticulitis, a radiological review of 95 cases demonstrated a higher incidence of jejunal involvement (58%) compared to duodenal involvement (26%) (6-7). Perforations involving the third and fourth portions of the duodenum are especially rare (2). Duodenal diverticula may be classified as either acquired extraluminal pseudodiverticula or congenital intraluminal diverticula, the latter occurring more commonly in pediatric populations (8). Given the different anatomic properties of the duodenum (e.g., as a primarily retroperitoneal organ), for purposes of the remainder of this review we focus on jejunal diverticulitis.

Patients with jejunal diverticulitis may be asymptomatic or present with nonspecific symptoms such as fever, nausea, vomiting, or jaundice. Abdominal manifestations can include bloating, discomfort, early satiety, and changes in bowel habits such as diarrhea or steatorrhea, sometimes related to bacterial overgrowth (9). Epigastric pain has also been reported (9-10). Complications of jejunal diverticulitis may include abscess formation, hemorrhage, perforation, fistula, volvulus, biliary obstruction, and bacterial thromboembolism (11). Among these, perforation is particularly concerning, carrying a mortality rate of up to 40% when diagnosis is delayed (12).

Cross-sectional imaging plays a central role in diagnosis. Both CT and MRI can aid in visualizing diverticula, though CT is widely regarded as the preferred modality (2,9). Mathis et al. recommend CT even in emergent situations, with sensitivity exceeding 90% (10,13). Other studies base imaging selection on symptomology, with ERCP, ultrasound, and EGD serving as adjunct modalities in select cases (13). Despite its utility, CT has limitations, as diverticula may be subtle and difficult to visualize, potentially resulting in delayed diagnosis and treatment (12).

Common contrast-enhanced CT findings described in the literature include contained perforation, peri-diverticular fat stranding, extraluminal gas, mesenteric edema, small fluid collections, and localized pneumoperitoneum (5,7). Non-contrast CT may also be helpful in diagnosing patients for whom contrast is contraindicated. A case series by Luna et al. (2025) used non-contrast CT and showed jejunal outpouching with mural discontinuity and surrounding mesenteric fat stranding, aiding in the diagnosis of a contained perforation (10). The differential for small bowel diverticular disease may include peptic ulcer disease, cholangitis, pancreatitis, colitis, and retrocecal appendicitis (8).

Management of jejunal diverticulitis depends largely on the patient’s clinical presentation and underlying comorbidities. Based on numerous case studies, management ranges from nonoperative management with antibiotics to urgent surgical intervention. Conservative management includes bowel rest, nasogastric decompression, total parenteral nutrition (TPN), broad-spectrum antibiotics, and, when indicated, percutaneous catheter drainage for source control (8). Non-operative management is generally reserved for patients who are hemodynamically stable or have significant contraindications to surgery (14). Intraoperative cultures and microbiological evaluation should be obtained when possible to support antibiotic stewardship (12).

While no specific classification for jejunal diverticulitis has been proposed, in terms of small bowel diverticulitis Chait et al. proposed a modified Hinchey classification to guide the categorization and management of duodenal diverticulitis (14). Endoscopic interventions are most often indicated for hemorrhagic complications or for diverticula located near the ampulla of Vater (14). For jejunal diverticulitis, whether persistent or intractable pain alone should prompt surgical intervention remains debated (13). Surgical options include open diverticulectomy, jejunal resection with side-to-side or end-to-end anastomosis, and laparoscopic approaches (12-13). Laparoscopy may be particularly appropriate for limited jejunal involvement and has been associated with reduced post-operative pain, morbidity, and length of stay (12).

In a retrospective case series of 12 patients with jejunal diverticulitis treated in the United Kingdom between 2012 and 2025, Desmay et al. reported a predominantly male cohort with a mean age in the early seventies, 7 of which underwent surgical intervention and the remainder managed non-operatively (4). Complication rates were comparable between groups (42.9% vs. 40%). Antibiotic regimens in this study followed institutional protocols derived from the National Institute for Health and Care Excellence (NICE) guidelines for complicated diverticular disease. Similarly, Santamaria et al. based antimicrobial therapy on established guidelines for community-acquired intra-abdominal infections without diffuse peritonitis or septic shock (12). Early detection and timely intervention remain critical to decrease mortality. Histopathologic evaluation may further assist in excluding malignancy and characterizing the underlying diverticular pathology (3,15).

Current evidence regarding jejunal diverticulitis is limited to case reports and small retrospective series, and there is no clear consensus on optimal operative timing for symptomatic small bowel diverticula (14). This study seeks to contribute to the existing literature by further characterizing management strategies and complication rates at our institution. Given that complication rates at our single academic center appear higher than those reported elsewhere, our goal is to better define treatment approaches that may ultimately reduce morbidity and mortality.

## Methods

We conducted a retrospective review of patients diagnosed with jejunal diverticulitis in a single academic center’s Emergency General Surgery registry between December 2017 and December 2024. Data abstracted included age, sex, imaging modality, presence of perforation, serial physical exams, lab values (CBC, lactate), ICU admission, length of stay (LOS), antibiotic duration, operative status and timing, distance of residence from our institution, disposition after index admission, and readmission within one year.

Continuous variables were summarized using means with standard deviations and ranges. Categorical variables were reported as counts and percentages. Comparative analyses between operative and non-operative patients were conducted using t-tests or Mann–Whitney U tests for continuous variables, and chi-square or Fisher’s exact tests for categorical variables. Statistical analyses were performed using IBM SPSS Statistics v. 31.0.0.0, with significance set at p < 0.05.

## Results

Figure 1 depicts the patient distribution from our review.

**Figure 1.**
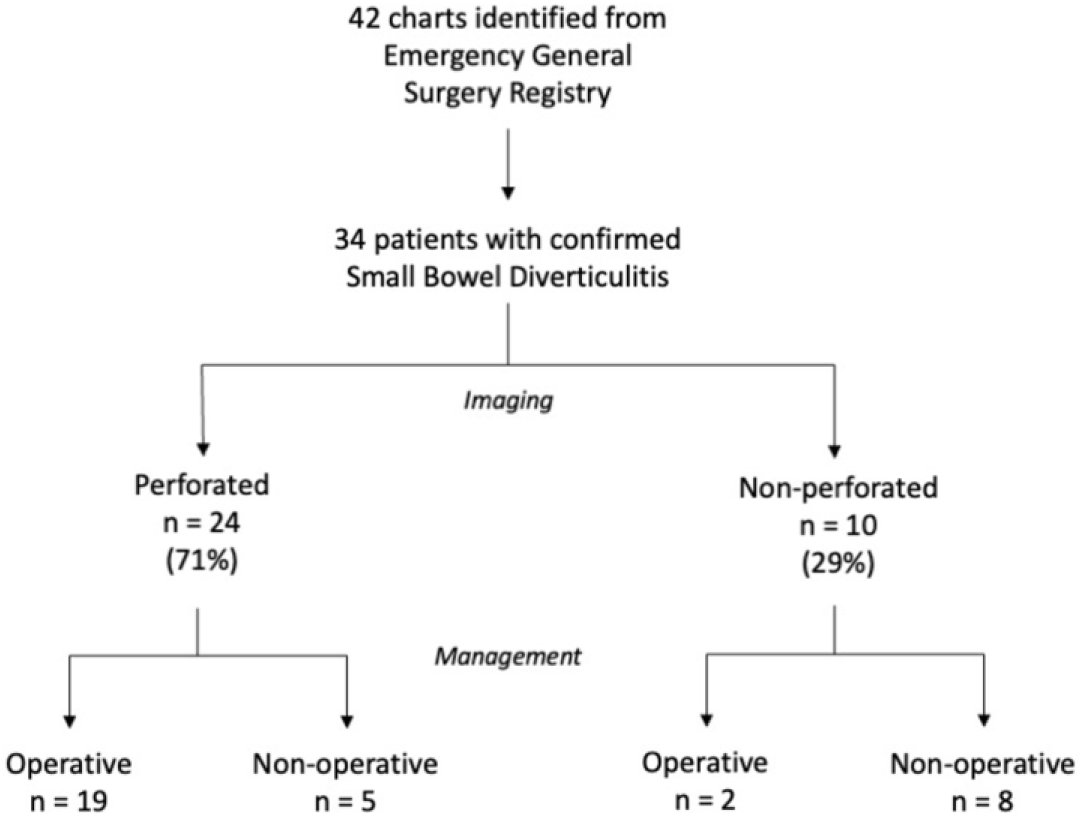
Review flow chart demonstrating distribution of patients in this study.

Table 1 shows the results of our review.

**Table 1.** Results of review.

|  | Total<br>(n) | Female<br>(n) | Male<br>(n) | Mean<br>Age | Age<br>Range | Leukocytosis<br>on Admission<br>(n) | Mean Duration<br>of Antibiotics<br>(days) | Mean<br>Length of<br>Stay (days) | ICU<br>Admission<br>(n) | Mean<br>Residence<br>Distance (mi) | Range<br>Residence<br>Distance (mi) |
| --- | --- | --- | --- | --- | --- | --- | --- | --- | --- | --- | --- |
| **Perforation** | 24 | 15 | 9 | 74.9 | 53-89 | 19 | 9.5 | 10.8 | 7 | 31.9 | 2-206 |
| Operative | 19 | 12 | 7 | 76.3 | 53-89 | 14 | 8 | 12 | 1 | 21.6 | 2-83 |
| Non-operative | 5 | 3 | 2 | 69.4 | 54-80 | 5 | 16.5 | 6.2 | 6 | 70.8 | 16-206 |
| **Non-perforated** | 10 | 4 | 6 | 74.4 | 68-91 | 8 | 14.7 | 7.3 | 1 | 18.6 | 5-62 |
| Operative | 2 | 1 | 1 | 70.0 | 68-72 | 2 | 22.5 | 11.5 | 0 | 53.5 | 45-62 |
| Non-operative | 8 | 3 | 5 | 75.5 | 68-91 | 6 | 12.7 | 6.3 | 1 | 9.9 | 5-20 |
| **Total** | 34 | 19 | 15 | 74.7 | 53-91 | 27 | 10.9 | 9.8 | 8 | 28.0 | 2-206 |

Of 42 patients initially identified with the diagnosis code of “small bowel diverticulitis” in the EGS Registry, 34 had confirmed small bowel diverticulitis on chart review and were included in the final analysis. The mean age was 74.7 years (SD 10.3; range 53–91). 19 patients (56%) were female and 15 (44%) were male.

The average distance from our medical center was 28.0 miles (SD 37.6; range 2–206). 19 patients (56%) were residents of Chittenden County. Distance and Chittenden County residence were not associated with perforation, ICU admission, or LOS.

All patients received antibiotic therapy. The mean duration of antibiotic treatment was 10.9 days (SD 7.9; range 4-21), with one outlier receiving 41 days of therapy. Duration of antibiotic therapy was significantly longer among patients with perforation compared to those without perforation (p = 0.037).

Twenty-four patients (71%) were diagnosed with perforation, representing a high-perforation cohort. There was no significant difference in age between perforated and non-perforated patients (74.9 ± 11.6 years vs. 74.4 ± 6.7 years). Leukocytosis was common in both groups and did not significantly differ by perforation status. Similarly, elevated lactate (>2 mmol/L), ICU admission, and Chittenden County residence were not significantly associated with perforation (all p > 0.05, Fisher’s exact test), although incomplete lactate data limited interpretation. 19 of these patients underwent operative management. Two of these patients presented with small bowel obstruction, 16 patients with abscess or contained perforations, and one patient with both abscess and contained perforation in two separate locations of the small bowel.

Seven patients (21%) required ICU admission, most of whom had perforation. The mean hospital length of stay (LOS) was 9.8 days (SD 6.8; range 2-30). Because LOS was non-normally distributed, a Mann-Whitney U test was performed, demonstrating a significant association between perforation and longer LOS (p = 0.015). The longest hospitalization (30 days) occurred in a perforated patient requiring ICU admission.

Two patients were readmitted within one year for reasons unrelated to their index admission. Three patients (9%) in this cohort died. Two deaths occurred during the index admission, both in perforated patients who underwent surgery. One additional patient died after readmission the day following discharge due to distributive shock secondary to decompensated cirrhosis.

### Operative Management

Overall, 21 patients (62%) underwent surgical intervention. Operative management was strongly associated with perforation: 19 of 24 perforated patients (79%) underwent surgery compared to 2 of 10 non-perforated patients (20%) (Fisher’s exact test, p = 0.002).

Nine patients proceeded directly to the operating room (five within one hour of arriving to the Emergency Department, four within 12 hours), three underwent surgery on hospital day 1, and two as late as hospital day 6. Five perforated patients were managed non-operatively.

All operations were performed via exploratory laparotomy. Small bowel resection was performed in 20 patients (95%), typically with primary anastomosis (90.5%). One patient underwent lysis of adhesions without resection. Two cases required staged abdominal operations with temporary abdominal closure following damage control laparotomies: one due to severely inflamed and friable tissue with concern for staple line integrity, and the other due to multiple areas of necrotic and hemorrhagic small bowel in the setting of hemodynamic instability requiring vasopressor support.

Pathologic evaluation consistently demonstrated diverticular disease with severe acute diverticulitis, frequently with transmural inflammation, perforation, abscess formation, acute serositis, and/or ischemic changes. No dysplasia or malignancy was identified in resected small bowel specimens, though one incidental appendiceal mucinous neoplasm was noted.

### Non-Operative Management

Ten patients (29%) were classified as non-perforated on index imaging. The mean age was 74.4 years (range 68-91) and 6 (60%) were male. Imaging demonstrated localized inflammatory changes without free intraperitoneal air or generalized peritonitis. A minority had small volume extraluminal gas or free fluid without organized collections. All patients were hemodynamically stable on presentation.

Five patients with CT-confirmed perforated jejunal diverticulitis were also managed non-operatively. These patients demonstrated contained perforation on imaging, hemodynamic stability, and absence of generalized peritonitis. Four of these patients (80%) underwent percutaneous image-guided drainage, while one was managed with antibiotics alone. Most demonstrated resolution of leukocytosis and improvement on serial abdominal examinations. One patient demonstrated rising leukocytosis and lactate during the hospital course, concerning for impending failure of non-operative management.

Compared to non-perforated patients, perforated patients were significantly more likely to require surgery (p = 0.002) and had longer average hospitalizations (10.8 days vs 7.8 days). There was a statistically significant difference in duration of antibiotic therapy between patients managed operatively and those managed non-operatively (p = 0.016, Mann-Whitney U test), with a shorter mean duration observed in the operative group (9.4 days) compared to the non-operative group (14.0 days). However, select perforated patients with contained disease were successfully managed non-operatively when clinically stable and closely monitored.

## Discussion

In this retrospective cohort of 34 patients with confirmed jejunal diverticulitis, we observed that perforation was common, occurring in 71% of cases, and was strongly associated with the need for operative management. Patients with perforated disease generally experienced longer hospital stays, though not statistically significant, and required longer courses of antibiotics, reflecting greater clinical severity and resource utilization. Notably, age, leukocytosis, elevated lactate, ICU admission, and geographic factors, including distance from the medical center or residence within Chittenden County, were not independently associated with perforation.

Our findings highlight the heterogeneity of jejnual diverticulitis and the potential for select perforated cases to be managed non-operatively. Five patients with CT-confirmed perforation were successfully treated without surgery. These patients were hemodynamically stable, lacked generalized peritonitis, and demonstrated contained perforations on imaging, often with abscess formation managed by percutaneous drainage. Serial laboratory monitoring and physical examinations demonstrated clinical improvement, underscoring the importance of careful patient selection and ongoing reassessment in non-operative management.

The mortality observed in this cohort (three patients, 9%) underscores the potential severity of jejunal diverticulitis, particularly among those with perforation and comorbid conditions. Importantly, two of the three deaths occurred in patients who underwent operative management, reflecting both disease severity and the complex interplay of underlying comorbidities, such as cirrhosis and immunocompromised status.

These results suggest several clinical implications. First, perforation alone should not dictate automatic surgical intervention; rather, patient hemodynamics, physiologic reserve, and radiologic features should guide management decisions. Second, serial clinical assessment and laboratory monitoring are critical in identifying candidates for non-operative care and ensuring timely escalation if deterioration occurs. Third, while geographic factors did not influence outcomes in our cohort, the mean distance from the hospital suggests that community and regional patients can safely undergo initial assessment and management at a tertiary center without apparent delays in care.

Overall, our study demonstrates that jejunal diverticulitis, though frequently presenting with perforation in an elderly cohort, can be managed effectively through individualized, risk-stratified approaches. Operative intervention remains necessary for many perforated patients, yet carefully selected non-operative strategies can achieve favorable outcomes while minimizing surgical morbidity. These findings contribute to the limited literature on the management of small bowel diverticulitis and may inform clinical decision-making in similar tertiary care settings.

This study has several limitations. First, its retrospective, single-center design introduces inherent risks of selection bias and limits generalizability. Jejunal diverticulitis, particularly with perforation, is a rare condition, and our modest sample size restricts statistical power and limits the ability to perform robust multivariable modeling. Additionally, missing laboratory data (notably lactate values) may have limited assessment of physiologic predictors of severity.

Second, the binary classification of perforation does not fully capture the spectrum of disease severity (e.g., contained vs. free perforation), which may influence management decisions and outcomes. Detailed radiographic characteristics and standardized severity grading were not consistently available. Finally, as management decisions were made at the discretion of treating surgeons, variability in operative thresholds and antibiotic duration may have influenced observed outcomes. Larger, multi-institutional studies with standardized definitions and management algorithms are needed to better characterize optimal treatment strategies for perforated small bowel diverticulitis.

## Conclusion

Compared to the 6% complication rate reported in prior literature, our series demonstrates a notably higher rate of perforation (71%) among patients diagnosed with jejunal diverticulitis. Operative intervention was common, though a subset of patients was successfully managed non-operatively with antibiotics. Mortality was limited to patients with significant comorbidities and complex presentations. These findings underscore the heterogeneity in presentation and outcomes, and highlight the need for a standardized approach. Development of practice guidelines incorporating clinical, radiographic, and laboratory parameters may improve diagnostic accuracy and guide timely, evidence-based management of this rare but serious condition.

## Data Availability

All data produced in the present study are available upon reasonable request to the authors.

